# Medicare Advantage Coupled with Dual Eligibility is Associated with Stroke Rehabilitation Outcomes Differences

**DOI:** 10.64898/2026.04.30.26352190

**Authors:** Amol M. Karmarkar, Charmi Kanani, Alexandra L. Terrill, Will Schroeder, Kimberly S. Erler, William E. Carter, Corey R. Fehnel, Amit Kumar

**Affiliations:** Department of Physical Medicine and Rehabilitation and Center for Rehabilitation Science and Engineering, Virginia Commonwealth University, Richmond, VA, USA; Clinical Science and Research Department, Sheltering Arms Institute, Richmond, VA, USA; Department of Physical Medicine & Rehabilitation, University of Utah, Salt Lake City, UT, USA; Department of Occupational & Recreational Therapies, University of Utah, Salt Lake City, UT, USA; Virginia Commonwealth University School of Medicine, Virginia Commonwealth University, Richmond, VA, USA; Department of Occupational Therapy, MGH Institute of Health Professions, Boston, MA, USA; Department of Neurology, Beth Israel Deaconess Medical Center, Harvard Medical School, Boston, MA, USA; and Hinda and Arthur Marcus Institute for Aging Research, Boston, MA, USA; Department of Physical Therapy & Athletic Training, University of Utah, Salt Lake City, UT, USA

**Keywords:** Stroke, Inpatient Rehabilitation Facilities, Medicare Advantage, Dual Eligible

## Abstract

**Importance:** Medicare–Medicaid dual eligible beneficiaries experience pronounced disparities in stroke recovery. However, it remains unclear whether inpatient rehabilitation services and outcomes are comparable between dual-eligible beneficiaries enrolled in Medicare fee-for-service (FFS) versus Medicare Advantage (MA) plans.

**Objective:** To compare rehabilitation therapy utilization and associated outcomes among dual-eligible beneficiaries enrolled in FFS versus MA plans with stroke.

**Design:** Retrospective cohort study.

**Setting:** Inpatient Rehabilitation Facilities (IRF).

**Participants:** Medicare beneficiaries admitted to IRF with stroke (*n*=125,782) between 2017 and 2019.

**Exposure:** Dual-eligible beneficiaries enrolled in FFS versus MA plans.

**Main Outcome Measures:** Total number of minutes of physical and occupational therapy provided within the first 2 weeks of IRF stay, self-care and mobility change scores, and 30-day all-cause hospital readmission.

**Results:** For the first 2 weeks of therapy utilization, we did not find significant differences between the four groups. Using the non-dual FFS beneficiaries and low category of change as a reference, we found significantly lower likelihood of achieving high change in self-care scores for the dual FFS (OR=0.73, 95% CI=0.69-0.76), and dual MA (OR=0.93, 95% CI=0.88-0.98). However, non-dual MA patients had a higher likelihood of changes in self-care scores (OR=1.17, 95% CI=1.13-1.22). Similar trends were found for the mobility change scores, compared to non-dual FFS: dual FFS (OR=0.72, 95% CI=0.68-0.75), and dual MA (OR=0.91, 95% CI=0.86-0.96) and non-dual MA (OR=1.16, 95% CI=1.12-1.20). For 30-day readmission risk, dual FFS showed a higher likelihood of readmission (OR=1.19, 95% CI=1.08-1.31), while non-dual MA had a significantly lower likelihood (OR=0.77, 95% CI=0.71-0.83).

**Conclusions and Relevance:** Although no differences in rehabilitation therapy utilization for stroke among dual-eligible beneficiaries, they had poorer functional recovery and higher 30-day readmission risk irrespective of FFS vs MA. Whereas non–dual-eligible MA beneficiaries experienced favorable outcomes. These findings underscore the importance of addressing post-IRF discharge needs among disadvantaged populations.

## Introduction

Stroke incidence remains very high. It is estimated that every 40 seconds someone in the U.S. has a stroke, affecting more than 795,000 people annually.^1^ Stroke survivors often experience functional deficits that requires postacute care in inpatient rehabilitation facilities (IRF) or skilled nursing facilities (SNF). Guidelines by the American Heart Association/American Stroke Association for postacute care include provision of rehabilitation services from IRF, SNF, Home Health (HH) services.^2^ Inpatient Rehabilitation Facilities (IRF) are either rehabilitation units within a hospital or freestanding facilities that provide physician-directed intensive rehabilitation that include at least three hours of combined rehabilitation therapy from occupational therapy (OT), physical therapy (PT), and speech language pathology (SLP) services.^2,3^ Stroke is also one of the 13 Centers for Medicare and Medicaid Services (CMS)-specified conditions, a regulatory requirement stating at least 60% IRF admissions must involve patients with one or more qualifying medical conditions.^3^

Medicare Advantage (MA) plans are the private and marketplace-based insurance alternative to traditional Medicare fee-for-service (FFS). There has been tremendous growth in the enrollment of individuals into MA plans, with recent data reporting more than 54% of Medicare beneficiaries are now enrolled in MA plans.^4^ This growth has implications for access to and utilization of health services, particularly postacute care. There are key differences in access to IRF services between FFS and MA plans. For example, IRF prospective system (IRF-PPS) which was initiated as part of the Balanced Budget Act of 1997 and implemented in 2002, uses the prospective determination of length of stay (LOS), using a case-mix grouping (CMG) system combined with comorbidities and other facility-level characteristics.^3,5^ In traditional FFS plans, ideal candidates for IRF admission are largely determined by the ability of the individual to tolerate intense rehabilitation services, and availability of social support that could increase the likelihood of discharge to the community after IRF stay.^2^ However MA plans differ from FFS in the following ways: 1) in addition to the ideal candidate determination there is a prior authorization process, that could delay early access to IRF, or denial of the IRF stay altogether.^6^ 2) MA plans utilize a limited network of facilities for patients to receive postacute care, and 3) manage access and utilization of postacute care through cost-sharing, and utilization management/review.^7–15^ Cumulatively, this limits access to IRF for MA beneficiaries, prior evidence also suggests differences in IRF utilization and outcomes among beneficiaries on Medicare FFS and MA plans, with MA beneficiaries demonstrating shorter LOS and better outcomes compared to FFS.^16–18^ However, these investigations were limited by use of registry data, and did not include the entire sample of Medicare beneficiaries on FFS and MA plans. Also, these studies did not include post-IRF outcomes, for example, 30-day hospital readmission.

The most pronounced disparities in recovery after stroke occur among adults covered by both Medicare and Medicaid (dual-eligible beneficiaries), a population that serves as a proxy for both socioeconomic disadvantage and high medical complexity.^19–21^ Concurrent with the expansion of MA plans, and the incorporation of Special Needs Plans (SNPs) into the MA program, the majority of dual-eligible beneficiaries are now enrolled in MA plans. Currently, approximately 8.3 million dual-eligible individuals (68%) are enrolled in MA, compared with 4.0 million (32%) in FFS Medicare.^22^ Dual-eligible beneficiaries disproportionately represent socio-economically and clinically vulnerable populations. Nearly half of dual-eligible beneficiaries are minorities, and 44% report fair or poor health, compared with 17% of non-dual eligible beneficiaries, and 48% report limitations in activities of daily living, compared with 23% of non-dual eligible beneficiaries.^23^ These disparities underscore the heightened need for effective postacute care among dual-eligible patients enrolled in MA plans following acute stroke. There is limited evidence to show differences in inpatient rehabilitation utilization among duals and non-duals eligible beneficiaries. To date only a single prior study has compared self-care and mobility outcomes between dual and non-dual beneficiaries and found no differences after adjusting for sociodemographic and socioeconomic status.^24^

Although a growing body of research has examined differences in outcomes among patients with stroke by Medicare insurance type, prior studies have largely examined FFS and MA populations separately, without accounting for the role of dual eligibility or examining differences by dual eligibility without accounting for MA status. The overlap between MA status and dual-eligibility is growing. Thus, examining the interaction between MA and FFS within the context of dual-eligibility status is critical. Additionally, there is no information on the sequential outcome differences between these populations in terms of: therapy utilization, functional outcomes, and 30-day hospital readmission across these populations. Addressing this gap is critical, as variation in postacute rehabilitation care following hospitalization for acute stroke may exacerbate existing disparities and represent modifiable opportunities to improve patient-centered outcomes. Although the Improving Medicare Post-Acute Transformation Act of 2014 aimed to increase transparency in therapy delivery and introduced standardized therapy-utilization reporting^25^, there remains a critical gap in understanding how therapy use varies across Medicare types, particularly for stroke which is one of the most resource-intensive IRF conditions. Therefore, the purpose of this study was to examine whether rehabilitation therapy utilization in IRF differs between dual-eligible beneficiaries admitted with stroke enrolled in FFS versus MA plans, and to explore associations with functional outcomes, and 30-day hospital readmission.

## Methods

### Ethics and Resource Sharing Statement

The study was approved by the institutional review board at Virginia Commonwealth University with a waiver of informed consent due to use of secondary deidentified data via a data use agreement (DUA) with the CMS. We followed the Strengthening the Reporting of Observational Studies in Epidemiology (STROBE) reporting guideline.^25^

#### Study Design and Data Sources

We utilized a 100% sample of patient-level Medicare claims data, including the Master Beneficiary Summary File (MBSF), Medicare Provider Analysis and Review (MedPAR) file, and postacute assessment files from January 1, 2017 to November 30, 2019. The MBSF contains information on beneficiaries’ sociodemographic characteristics, and indicators for Medicare enrollment such as MA, FFS, and dual eligible status. The MedPAR contains all claims information related to acute hospitalization. The Inpatient Rehabilitation Facility Patient Assessment Instrument (IRF-PAI) contains detailed direct patient assessments for all IRF admissions and discharges, regardless of the patient’s insurance status (MA and FFS).

#### Study Sample Population

The cohort included Medicare beneficiaries aged 65 and older enrolled either in FFS or MA plans, who were admitted to acute hospitals between January 1, 2017, and discharged from IRF on/before November 30, 2019, with ischemic stroke. The diagnosis of ischemic stroke was identified using the primary International Classification of Diseases, Tenth Revision, Clinical Modification (ICD-10 CM) codes (I63.x and I67.8). We included patients discharged from acute care hospitals, followed by direct admission (operationally defined as within 48 hours) to IRF. To capture valid IRF stays, admission to IRF was determined using the subsequent claims and assessments information from IRF-PAI. We also excluded patients who had an acute hospitalization LOS exceeding 30 days, were admitted to the hospital from SNF or an Intermediate Care Facility or were discharged to a hospice or long-term care facility (nursing homes). The final sample consisted of 125,782 patients with ischemic stroke who were discharged to IRF after an acute hospitalization (Figure 1).

**Figure 1.**
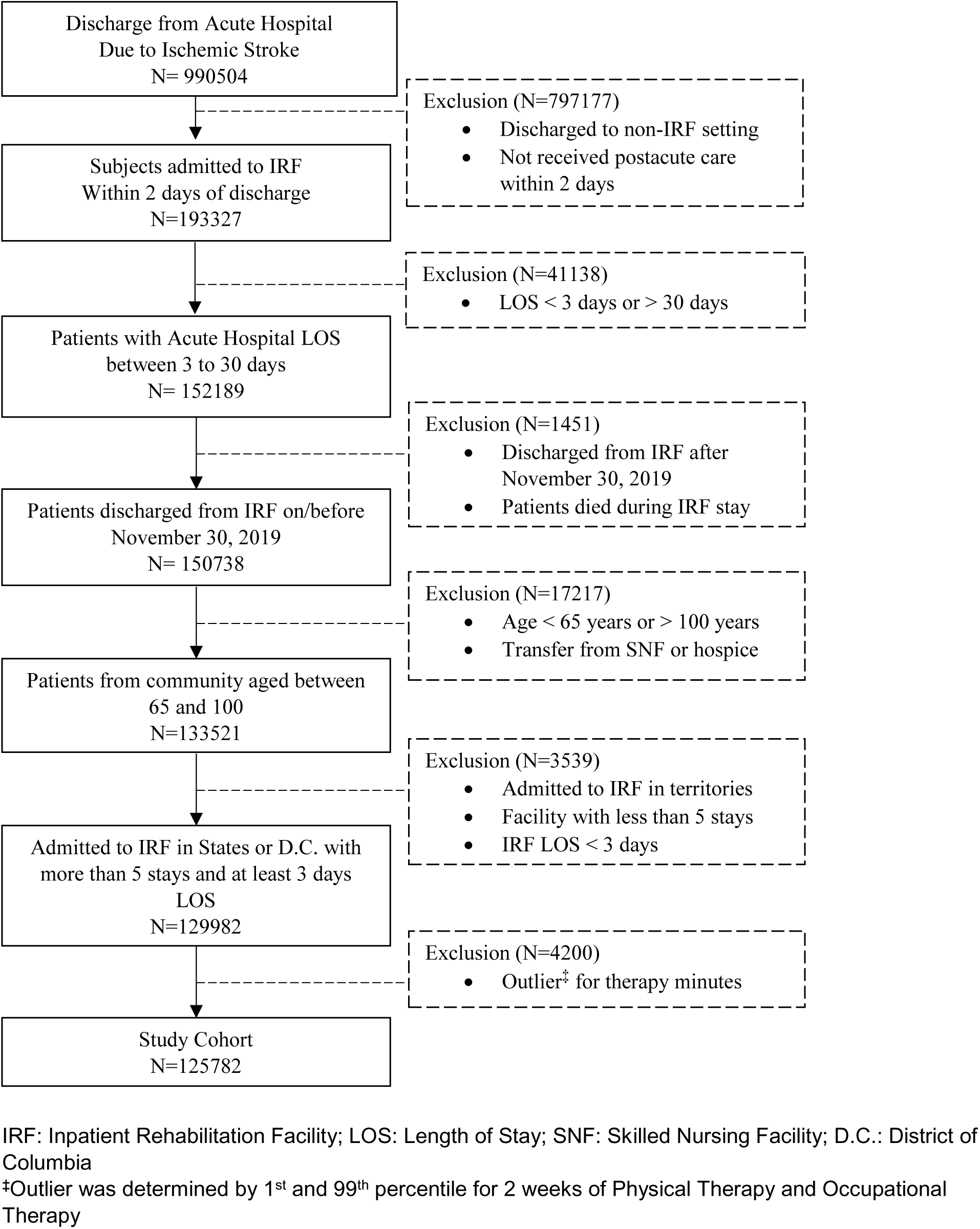
Flowchart of Cohort Selection. IRF: Inpatient Rehabilitation Facility; LOS: Length of Stay; SNF: Skilled Nursing Facility; D.C.: District of Columbia ^‡^Outlier was determined by 1^st^ and 99^th^ percentile for 2 weeks of Physical Therapy and Occupational Therapy

#### Primary Variable of Interest

The primary independent variables were 1) MA versus FFS; and 2) dual-eligible status (dual vs non-dual), which were obtained from the MBSF. The MBSF contains data on beneficiary characteristics and monthly enrollment in Part A (inpatient), Part B (outpatient), Part C (Medicare Advantage/ managed care/ HMO) and Part D (prescription drug coverage) of all Medicare beneficiaries enrolled in Medicare within a given calendar year. Beneficiaries were classified as MA or FFS based on their enrollment status in the MBSF at the time of discharge from the stroke hospitalization. Dual-eligible patients were classified if they were enrolled in both Medicare and Medicaid as determined by their enrollment status in the MBSF, at the time of discharge from the stroke hospitalization and during their IRF stay. We created four mutually exclusive categories: non-dual FFS, dual FFS, non-dual MA, and dual MA.

#### Study Variables

##### Rehabilitation Therapy Minutes

Rehabilitation therapy minutes were defined as the total number of minutes provided by physical therapy and occupational therapy within the first 2 weeks in IRF stay. A week was defined as a 7 consecutive calendar day period starting with the day of admission. CMS requires standardized reporting of patients’ therapy time through the IRF-PAI Therapy Information Section (Items O0401 and O0402), but these data elements are collected for reporting rather than to determine whether IRFs meet coverage requirements.^12,13^ Extreme values (outliers) outside of 1% at the minimum and 99% at the maximum weekly therapeutic time were removed (n=2461). The tertile (low, medium, and high) of OT, PT, and total treatment minutes was based on the distribution of those categories. The total number of minutes was used to estimate variation across facilities, and the tertile of total treatment time was used to estimate the influence of patients’ characteristics and facility-level factors. Speech-language pathology (SLP) minutes were not included in the current study. We also calculated a combined therapy utilization (OT plus PT) to examine the interaction between the four groups and therapy utilization on self-care and mobility change scores.

##### Self-Care and Mobility Change

Using previously published methods, section GG within IRF-PAI assessment were used to calculate admission, discharge, and change scores for self-care and mobility domains.^24,26,27^ We also calculated overall functional status change (self-care plus mobility score) to test the interaction effect between the four groups with the overall functional status change and whether it is associated with 30-day readmission.

##### All Cause 30-Day Hospital Readmission

We calculated the 30-day all-cause risk-adjusted hospital readmission (yes/no) for each patient after discharge from IRF. We did not include patients who were transferred to acute hospitals at the end of their IRF stay (n=6,868). Therefore, we used a smaller cohort of individuals for the readmission model (n=118,914). We used 30-day mortality as a competing event for estimating the risk of 30-day all cause hospital readmission.^28^

##### Patient-Level Covariates

Demographic and clinical severity measures were accounted for in the analysis. Demographic characteristics included age groups (65-74, 75-84, ≥85), sex (male/female), and race/ethnicity (non-Hispanic white, non-Hispanic Black, Hispanic, and other). These variables were extracted from the MBSF file. Clinical severity factors included length of stay in IRF, Elixhauser comorbidity (<3 and ≥3), and CMS tier comorbidity levels (0,1,2,3). Rehabilitation LOS was total number of days in the IRF. We utilized 25 ICD-10 codes from the comorbid condition section of the IRF-PAI to create an Elixhauser Comorbidity score.^9^ Using previously published methods, ICD-10 diagnosis codes were used to calculate the sum of each patient’s comorbid conditions based on 31 medical conditions in the Elixhauser Comorbidity index.^29^ To align with CMS payment policy, we adjusted the Tier comorbidity algorithm developed by CMS following the implementation of the prospective payment system in IRF. This system classifies medical conditions into four categories: no tiered comorbidity, Tier 1, Tier 2, and Tier 3, based on the costs incurred during an inpatient rehabilitation stay. Tier 1 represents the highest cost category and includes eight diagnostic codes, while Tier 3 encompasses 932 diagnostic codes and is the lowest cost tier. The IRF-PAI file contains information on the Tier categories for each patient.^7^ Detailed information related to all the patient-level variables is provided in eTable 6.

##### Facility-Level Covariates

We linked the data with IRF Rate Setting Files to classify facilities as freestanding or rehabilitation units, rurality of the facility (urban/rural), profit status (for profit/not-for-profit), safety net status (yes/no), and the volume of each facility using the number of total discharges (overall volume).^30^ The total number of MA and dual-eligible beneficiaries in each facility were classified in tertiles (low, medium, high).

##### Regional Characteristics

All-cause readmission risk adjusted for regional factors for each patient. We used the Robert Wood Johnson Foundation (RWJF) County Rating File and looked at county ranking for: clinical care, socio-economic factors, and health outcomes.^31^ The RWJF ranking system uses a quartile-based ranking (1-4). We used this to further classify counties in high (1) and low (2-4) for the three factors mentioned. In addition, CMS data were used to determine the penetration rate of MA plans in a county. MA penetration is calculated by dividing the number of beneficiaries enrolled in MA plans by the number of eligible Medicare beneficiaries in a county for a given month.^32^ We used the median value of average MA penetration and classified the counties into low and high penetration.

## Data Analysis

Demographic and clinical factors were examined descriptively (mean and percentage) by the four groups using Medicare insurance type (FFS vs MA) and dual eligibility status. Baseline characteristics and individual therapy time were compared with ANOVA or Chi-square test. For OT and PT utilization, we used a generalized linear mixed model (GLMM) with multinomial distribution and glogit link to model utilization of rehabilitation therapy accounting for patients nested in IRF, adjusting for patient-level covariates (sociodemographic, LOS, comorbidities), and facility-level characteristics. To model self-care and mobility score change (low, medium, high), we used GLMM with multinomial distribution and glogit link, accounting for patient- and facility-level characteristics. To assess the effect of rehabilitation therapy utilization on self-care and mobility score change, we tested the interaction effect of overall rehabilitation therapy utilization (combined OT and PT) with the four mutually exclusive groups. Finally, we modeled the adjusted risk of 30-day all-cause readmission using mortality as a competing event, because patients who die cannot experience a hospital readmission. We used competing risk analysis, a specialized form of time-to-event analysis, in the presence of a competing event (30-day mortality), adjusting for patient-level covariates. In addition to using IRF-level characteristics for the 30-day readmission risk, we also used county rankings in the areas of: clinical care, socio-economic factors, and health outcomes. Similar to functional change, for 30-day readmission, we tested the interaction effect of the combined functional change score with the four mutually exclusive groups and then conducted post-hoc analysis stratified analysis by functional change score (categories) for the cox hazard models, using the Gray’s test.^33^ All analyses were conducted using Python and SAS 9.4 (SAS Inc., Cary, NC).^a^

## Results

### Medicare Insurance Plan Types (FFS vs MA) and Dual Eligibility

Our study cohort consisted of 125,782 Medicare beneficiaries with stroke admitted to IRFs. There were 70,291 (55.88%) individuals in non-dual FFS, 14,479 (11.51%) dual FFS, 31,235 (24.83%) non-dual MA, and 9,777 (7.77%) dual MA. We found significant differences in sociodemographic characteristics, comorbidities, health outcomes, facility-level factors between the four groups. (Table 1). For 30-day hospital readmission, after further excluding individuals who were discharged to acute hospitals (n=118,914), we found significant differences between the groups by regional characteristics (eTable 1).

**Table 1.** Descriptive Group Differences by Patient and Facility Characteristics-Overall (N = 125,782)

| Variable | Total | FFS Non-Dual | FFS Dual | MA Non-Dual | MA Dual |
| --- | --- | --- | --- | --- | --- |
| Patient-Level |  |  |  |  |  |
| Number of Subject | 125782 | 70291 | 14479 | 31235 | 9777 |
| Age, years, n(%)<sup>*</sup> |  |  |  |  |  |
| 65-74 | 49826 (39.6) | 25419 (36.2) | 6851 (47.3) | 12436 (39.8) | 5120 (52.4) |
| 75-84 | 49106 (39.0) | 27813 (39.6) | 5087 (35.1) | 12777 (40.9) | 3429 (35.1) |
| ≥ 85 | 26850 (21.3) | 17059 (24.3) | 2541 (17.5) | 6022 (19.3) | 1228 (12.6) |
| Sex, n(%)<sup>*</sup> |  |  |  |  |  |
| Male | 60310 (47.9) | 34916 (49.7) | 5835 (40.3) | 15666 (50.2) | 3893 (39.8) |
| Female | 65472 (52.1) | 35375 (50.3) | 8644 (59.7) | 15569 (49.8) | 5884 (60.2) |
| Race/Ethnicity, n(%)<sup>*</sup> |  |  |  |  |  |
| Non-Hispanic White | 93395 (74.3) | 57851 (82.3) | 7652 (52.8) | 23619 (75.6) | 4273 (43.7) |
| Non-Hispanic Black | 18836 (15.0) | 7406 (10.5) | 3441 (23.8) | 4786 (15.3) | 3203 (32.8) |
| Hispanics | 8010 (6.4) | 2594 (3.7) | 2077 (14.3) | 1714 (5.5) | 1625 (16.6) |
| Others | 4513 (3.6) | 1891 (2.7) | 1173 (8.1) | 865 (2.8) | 584 (6.0) |
| Unknown | 1028 (0.8) | 549 (0.8) | 136 (0.9) | 251 (0.8) | 92 (0.9) |
| Elixhauser Score, n(%)<sup>*</sup> |  |  |  |  |  |
| < 3 | 14909 (11.9) | 8386 (11.9) | 1531 (10.6) | 3883 (12.4) | 1109 (11.3) |
| ≥ 3 | 110873 (88.1) | 61905 (88.1) | 12948 (89.4) | 27352 (87.6) | 8668 (88.7) |
| Tier, n(%)<sup>*</sup> |  |  |  |  |  |
| 0 | 54701 (43.5) | 30575 (43.5) | 5297 (36.6) | 14875 (47.6) | 3954 (40.4) |
| 1 | 1465 (1.2) | 854 (1.2) | 268 (1.9) | 246 (0.8) | 97 (1.0) |
| 2 | 37257 (29.6) | 20607 (29.3) | 5232 (36.1) | 8257 (26.4) | 3161 (32.3) |
| 3 | 32359 (25.7) | 18255 (26.0) | 3682 (25.4) | 7857 (25.2) | 2565 (26.2) |
| LOS in IRF, mean (SD)<sup>*</sup> | 15.4 (7.02) | 15.2 (6.78) | 15.9 (6.92) | 15.3 (7.29) | 16.0 (7.90) |
| Physical Therapy<sup>*</sup> |  |  |  |  |  |
| Mean (SD) | 601 (207) | 597 (206) | 611 (206) | 600 (210) | 609 (206) |
| Low | 42568 (33.8) | 24064 (34.2) | 4539 (31.3) | 10778 (34.5) | 3187 (32.6) |
| Medium | 43232 (34.4) | 24374 (34.7) | 5140 (35.5) | 10382 (33.2) | 3336 (34.1) |
| High | 39982 (31.8) | 21853 (31.1) | 4800 (33.2) | 10075 (32.3) | 3254 (33.3) |
| Occupational Therapy<sup>*</sup> |  |  |  |  |  |
| Mean (SD) | 610 (199) | 607 (198) | 618 (199) | 609 (202) | 621 (198) |
| Low | 42085 (33.5) | 23857 (33.9) | 4516 (31.2) | 10667 (34.2) | 3045 (31.1) |
| Medium | 42841<br>(34.1) | 23987<br>(34.1) | 5088 (35.1) | 10372<br>(33.2) | 3394 (34.7) |
| High | 40856<br>(32.5) | 22447<br>(31.9) | 4875 (33.7) | 10196<br>(32.6) | 3338 (34.1) |
| Total Therapy* |  |  |  |  |  |
| Mean (SD) | 1210 (383) | 1200 (381) | 1230 (381) | 1210 (389) | 1230 (380) |
| Low | 42127<br>(33.5) | 23799<br>(33.9) | 4518 (31.2) | 10700<br>(34.3) | 3110 (31.8) |
| Medium | 41743<br>(33.2) | 23585<br>(33.6) | 4897 (33.8) | 10002<br>(32.0) | 3259 (33.3) |
| High | 41912<br>(33.3) | 22907<br>(32.6) | 5064 (35.0) | 10533<br>(33.7) | 3408 (34.9) |
| Self-care Change Score* |  |  |  |  |  |
| Mean (SD) | 9.39 (7.65) | 9.37 (7.68) | 8.61 (7.53) | 9.83 (7.62) | 9.25 (7.68) |
| Low | 46584<br>(37.0) | 26022<br>(37.0) | 6220 (43.0) | 10569<br>(33.8) | 3773 (38.6) |
| Medium | 42844<br>(34.1) | 24001<br>(34.1) | 4627 (32.0) | 10996<br>(35.2) | 3220 (32.9) |
| High | 36354<br>(28.9) | 20268<br>(28.8) | 3632 (25.1) | 9670 (31.0) | 2784 (28.5) |
| Mobility Change Score* |  |  |  |  |  |
| Mean (SD) | 25.4 (20.2) | 25.4 (20.2) | 22.6 (20.4) | 26.7 (19.9) | 24.8 (20.4) |
| Low | 43543<br>(34.6) | 24255<br>(34.5) | 5841 (40.3) | 9959 (31.9) | 3488 (35.7) |
| Medium | 42017<br>(33.4) | 23396<br>(33.3) | 4731 (32.7) | 10582<br>(33.9) | 3308 (33.8) |
| High | 40222<br>(32.0) | 22640<br>(32.2) | 3907 (27.0) | 10694<br>(34.2) | 2981 (30.5) |
| Functional Change Score* |  |  |  |  |  |
| Mean (SD) | 34.8 (25.7) | 34.8 (25.7) | 31.2 (25.7) | 36.5 (25.4) | 34.0 (25.8) |
| Low | 42112<br>(33.5) | 23547<br>(33.5) | 5706 (39.4) | 9469 (30.3) | 3390 (34.7) |
| Medium | 42890<br>(34.1) | 23783<br>(33.8) | 4874 (33.7) | 10870<br>(34.8) | 3363 (34.4) |
| High | 40780<br>(32.4) | 22961<br>(32.7) | 3899 (26.9) | 10896<br>(34.9) | 3024 (30.9) |
| Facility-Level |  |  |  |  |  |
| MA Beneficiaries, n(%)* |  |  |  |  |  |
| Low | 15715<br>(12.5) | 10300<br>(14.7) | 2790 (19.3) | 1902 (6.1) | 723 (7.4) |
| Medium | 33824<br>(26.9) | 19929<br>(28.4) | 4270 (29.5) | 7336 (23.5) | 2289 (23.4) |
| High | 76243<br>(60.6) | 40062<br>(57.0) | 7419 (51.2) | 21997<br>(70.4) | 6765 (69.2) |
| Dual Eligible Beneficiaries, n(%)* |  |  |  |  |  |
| Low | 16675<br>(13.3) | 10425<br>(14.8) | 1478 (10.2) | 3978 (12.7) | 794 (8.1) |
| Medium | 35643<br>(28.3) | 20640<br>(29.4) | 3780 (26.1) | 8965 (28.7) | 2258 (23.1) |
| High | 73464<br>(58.4) | 39226<br>(55.8) | 9221 (63.7) | 18292<br>(58.6) | 6725 (68.8) |
| Number of Discharges, mean (SD)* | 559 (486) | 579 (491) | 567 (518) | 519 (457) | 534 (479) |
| Rurality, n(%)* |  |  |  |  |  |
| Urban | 118953<br>(94.6) | 66030<br>(93.9) | 13465<br>(93.0) | 30075<br>(96.3) | 9383 (96.0) |
| Rural | 6829 (5.4) | 4261 (6.1) | 1014 (7.0) | 1160 (3.7) | 394 (4.0) |
| Safety-net Facility, n(%)* |  |  |  |  |  |
| Yes | 31801<br>(25.3) | 15501<br>(22.1) | 4948 (34.2) | 7882 (25.2) | 3470 (35.5) |
| No | 93981<br>(74.7) | 54790<br>(77.9) | 9531 (65.8) | 23353<br>(74.8) | 6307 (64.5) |
| Facility Ownership, n(%)* |  |  |  |  |  |
| Non-profit/Government | 79688<br>(63.4) | 43797<br>(62.3) | 9050 (62.5) | 20594<br>(65.9) | 6247 (63.9) |
| Profit | 46094<br>(36.6) | 26494<br>(37.7) | 5429 (37.5) | 10641<br>(34.1) | 3530 (36.1) |
| Freestanding Facility, n(%)* |  |  |  |  |  |
| Yes | 48651<br>(38.7) | 28227<br>(40.2) | 5407 (37.3) | 11413<br>(36.5) | 3604 (36.9) |
| No | 77131<br>(61.3) | 42064<br>(59.8) | 9072 (62.7) | 19822<br>(63.5) | 6173 (63.1) |

\* p<0.05
MA: Medicare Advantage; IRF: Inpatient Rehabilitation Facility; LOS: Length of Stay; FFS: Fee-for-Service

### Therapy Utilization

Table 2 and eTable 2 provide comparisons between the four groups adjusting for patient-, and facility-level variables on OT and PT utilization. Using non-dual FFS as the reference group, we did not find significant differences for occupational therapy utilization. For physical therapy utilization, compared to non-dual FFS group, we only found lower likelihood of receiving medium utilization of physical therapy for the non-dual MA group (OR=0.96, 95% CI=0.92-0.99).

**Table 2.** Association of Medicare Plan Type and Dual-Eligible Status on Inpatient Rehabilitation Outcomes.

| Variable | Physical Therapy |  | Occupational Therapy |  | Self-care Change Score |  | Mobility Change Score |  |
| --- | --- | --- | --- | --- | --- | --- | --- | --- |
|  | Medium vs Low | High vs Low | Medium vs Low | High vs Low | Medium vs Low | High vs Low | Medium vs Low | High vs Low |
|  | OR (95% CI) | OR (95% CI) | OR (95% CI) | OR (95% CI) | OR (95% CI) | OR (95% CI) | OR (95% CI) | OR (95% CI) |
| **Groups** |  |  |  |  |  |  |  |  |
| FFS Non-dual | Reference |  |  |  |  |  |  |  |
| FFS Dual | 1.01 (0.96-1.06) | 0.99 (0.94-1.04) | 1.00 (0.95-1.05) | 0.99 (0.94-1.05) | **0.80 (0.77-0.84)** | **0.73 (0.69-0.76)** | **0.84 (0.81-0.88)** | **0.72 (0.68-0.75)** |
| MA Non-dual | **0.96 (0.92-0.99)** | 0.97 (0.93-1.01) | 0.97 (0.94-1.01) | 0.97 (0.93-1.01) | **1.11 (1.07-1.14)** | **1.17 (1.13-1.22)** | **1.10 (1.06-1.14)** | **1.16 (1.12-1.20)** |
| MA Dual | 0.96 (0.91-1.02) | 0.95 (0.89-1.02) | 1.00 (0.94-1.06) | 0.99 (0.93-1.05) | **0.91 (0.86-0.96)** | **0.93 (0.88-0.98)** | 1.00 (0.94-1.05) | **0.91 (0.86-0.96)** |
| **Total Rehabilitation Therapy** |  |  |  |  |  |  |  |  |
| Low | NA | NA | NA | NA | Ref | Ref | Ref | Ref |
| Medium | NA | NA | NA | NA | **1.13 (1.08-1.18)** | **1.34 (1.28-1.41)** | **1.21 (1.16-1.27)** | **1.23 (1.17-1.29)** |
| High | NA | NA | NA | NA | **1.28 (1.22-1.35)** | **1.86 (1.76-1.96)** | **1.49 (1.42-1.57)** | **1.64 (1.55-1.73)** |
| **Interaction, joint test** |  |  |  |  |  |  |  |  |
| Plan*Therapy | NA | NA | NA | NA | **p=0.0002\*** |  | p=0.67 |  |

\* p<0.05
MA: Medicare Advantage; FFS: Fee-for-Service

### Functional outcomes

Using the non-dual FFS and the low-change category as the reference group, dual FFS group (OR = 0.73, 95% CI = 0.69-0.76), and dual MA group (OR = 0.93, 95% CI = 0.88-0.98) had reduced odds of achieving high change in self-care scores. In contrast, the non-dual MA group exhibited higher likelihood of achieving high change in self-care scores (OR = 1.17, 95% CI = 1.13-1.22). Similar trends were found for changes in mobility scores (Table 2 and eTable 2). Compared to non–dual FFS groups, the likelihood of change in high mobility score was lower among dual FFS (OR= 0.72, 95% CI= 0.68–0.75) and dual MA groups (OR= 0.91, 95% CI= 0.86–0.96). Non-dual MA group exhibited higher likelihood of achieving high change in mobility scores (OR=1.16, 95% CI=1.12-1.20).

We also tested the interaction between the four groups and overall therapy utilization and found significant interaction with the self-care change score (p=0.0002), but not with the mobility change score (p=0.6738) (Table 2 and eTable 2).

#### 30-day Readmission

For 30-day readmission risk, relative to non-dual FFS, we found a higher likelihood of readmission risk for dual FFS group (OR=1.19, 95% CI=1.08-1.31), but a lower likelihood for non-dual MA group (OR=0.77, 95% CI=0.71-0.83). We found a significant interaction between the overall functional status change and the four groups regarding the 30-day readmission (p=0.002) (Table 3 and eTable 5). Therefore, we performed a post-hoc analysis and presented CIF stratified by the three levels of functional change (low, medium, and high). Figure 2 delineates results stratified by all three functional change categories with significant differences in 30-day readmission rate for the four groups (p<.0001).

**Table 3.** Association of Medicare Plan Type and Dual-Eligible Status on 30-day Hospital Readmission.

| Variable | 30-Day Readmission |
| --- | --- |
|  | HR (95% CI) |
| **Groups** |  |
| FFS Non-dual | Ref |
| FFS Dual | **1.19(1.08-1.31)** |
| MA Non-dual | **0.77(0.71-0.83)** |
| MA Dual | 0.98(0.87-1.11) |
| **Total Functional Change** |  |
| Low | **1.75(1.66-1.85)** |
| Medium | **1.16(1.10-1.23)** |
| High | Ref |
| **Interaction, joint test** |  |
| Plan*Functional Change | **p=0.002\*** |

\* p<0.05
MA: Medicare Advantage; FFS: Fee-for-Service

**Figure 2.**
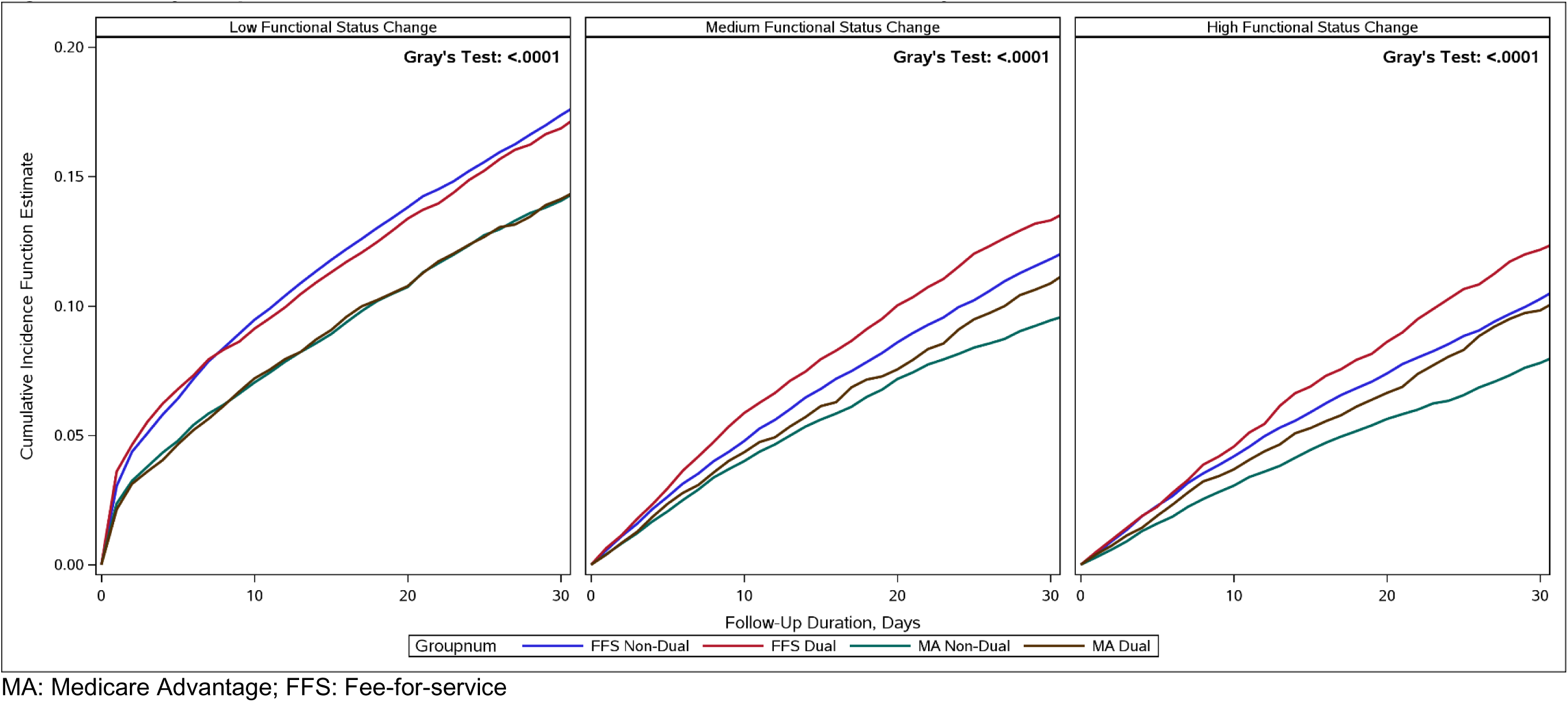
30-day Hospital Readmission Cumulative Incidence Function Stratified by Functional Status. MA: Medicare Advantage; FFS: Fee-for-service

## Discussion

In this nationally representative study of Medicare beneficiaries with stroke admitted to IRFs, rehabilitation therapy minutes did not differ significantly between FFS and MA status among dual-eligible beneficiaries. However, significant differences in functional outcomes and 30-day hospital readmission emerged by dual-eligibility status across FFS and MA plans. Dual-eligible beneficiaries in both FFS and MA plans had lower odds of achieving high self-care and mobility improvement, whereas non-dual MA beneficiaries were more likely to achieve improvement. A similar directional pattern was observed for 30-day readmissions, with higher risk among dual eligible FFS beneficiaries and lower risk among non-dual eligible MA beneficiaries. These findings highlight meaningful differences in recovery trajectories across Medicare enrollment groups, even when therapy utilization appears comparable.

Our findings align with previous findings limited to FFS and MA patients, excluding the dual eligibility population. A study of IRF admissions after stroke revealed a shorter LOS (1.5%) and greater likelihood of discharge to the community (3.0%) for MA patients compared to FFS patients with no significant difference in functional outcomes.^4^ Another recently published study compared risk-adjusted self-care and mobility scores among patients with and without dual eligibility status admitted to IRF and found slightly lower risk-adjusted improvements for self-care and mobility scores for dual eligible compared to non-dual eligible patients. Although the authors concluded that these differences were not clinically meaningful.^24^ Together, findings from our study extends understanding of differential patient outcomes and highlight dual-eligibility status as an important factor underlying persistent disparities after stroke.

To contextualize these findings, it is important to consider how therapy delivery is structured within IRFs. IRFs are expected to meet the longstanding intensity standard of providing at least 3 hours of therapy per day (or 15 hours per week), which facilities may implement in different ways.^13^ Patient characteristics such as age, medical complexity, and LOS, as well as facility characteristics, were also associated with therapy minutes, indicating that multiple factors shape therapy delivery beyond insurance enrollment alone. Using newly available patient-level function and outcomes data, this study adds novel evidence by demonstrating marked differences in recovery and rehospitalization across Medicare enrollment groups, even in the context of similar therapy utilization after stroke. Dual-eligible beneficiaries in MA and FFS plans had consistently lower functional gains and higher readmission risk, whereas non-dual MA beneficiaries demonstrated more favorable outcomes overall. These findings align with prior literature identifying dual eligibility as a marker of medical, social, and economic vulnerability that can influence recovery trajectories after stroke, independent of the care delivered during the IRF stay. Importantly, such differences carry particular significance for stroke, a condition characterized by complex motor, cognitive, communicative, and emotional sequelae that make recovery especially sensitive to social resources, caregiver support, and care coordination, which may be more difficult to access for dual-eligible beneficiaries. For non-dual eligible MA beneficiaries, enhanced care coordination or differences in baseline health status may contribute to the more favorable outcomes observed. Taken together, these results suggest that factors beyond therapy dose, including social risk, health complexity, and post-discharge resources, may shape functional recovery and rehospitalization risk for stroke survivors. Recent literature suggests a significant role of Social Determinants of Health (SDoH), which account for more than 50% of health outcomes, whereas clinical care accounts for only 20% of outcomes.^34^ In the context of inpatient rehabilitation facilities where the ultimate goal is discharge to the community, examining the role of SDoH, particularly, transportation availability, housing stability, and social support is important.

Stroke rehabilitation in IRFs is resource-intensive and high stakes, occurring during a critical window of neuroplasticity in which early gains can shape long-term recovery. Understanding how different factors contribute to recovery is increasingly important as Medicare continues to shift toward value-based models. The absence of differences in therapy utilization across Medicare groups, alongside clear variation in functional outcomes and readmissions, suggests that recovery after stroke is shaped by influences beyond therapy minutes alone. Clarifying the patient-, facility-, and system-level factors that contribute to these patterns will be important for improving the consistency and effectiveness of IRF stroke care. Future research should focus on identifying which aspects of IRF care and Medicare policy best support functional gains and reduce avoidable readmissions for all beneficiaries.

Prior investigations have demonstrated functional scores as a potential modifiable factor to mitigate risk for hospital readmission.^35–37^ In this study, we examined and found combined self-care and mobility changes and as a risk factor for 30-day hospital readmission, especially those in the low functional status change category. We also demonstrated differentiating risk regarding functional status change categories (low, medium, and high) among the four groups.

## Limitations

There are several important limitations to our study. First, the limitations associated with use of Medicare administrative claims and assessment data for research purposes are well documented. For our study, we only examined minutes for OT and PT services for the first two weeks of stay in IRF to estimate therapy utilization. The first two weeks are crucial during the early post-stroke recovery period; however, rehabilitation services and functional recovery can span several months, or even years, following stroke, which were not captured by this analysis. We did not have access to information on what specific types of rehabilitation interventions were provided or more granular information related to the therapy utilization. In the future, replicating some of this work using the compiled Electronic Health Records (EHR) from several IRF and classifying types of rehabilitation interventions that are being provided would be important. Second, while we risk-adjusted for comorbidities, we did not have access to stroke severity variables, neither we compare the findings by stroke sub-types. Finally, we assigned insurance type group membership (FFS vs MA and dual vs non-dual eligible) at the start of IRF admission and did not account for individuals switching from MA to FFS or vice-versa during the study duration. Finally, this study is observational and cannot establish causal relationships. Prior research has suggested selection bias in MA enrollment, and unobserved differences between FFS and MA beneficiaries may not be fully captured in administrative data, potentially influencing the observed associations. Despite these limitations, this study advances understanding of how rehabilitation therapy utilization and outcomes differ by Medicare coverage type and dual-eligibility status, contributing new evidence to an area with limited prior research. Additionally, this study leverages newly available patient-level functional data (section GG) among MA beneficiaries that have not been used previously, thereby strengthening the robustness of risk adjustment and enhancing relevance to clinical practice.

## Conclusion

With continued growth in MA enrollment and increasing beneficiary participation in Special Needs Plans since 2018, this study highlights concerning outcome disparities faced by dual-eligible beneficiaries. These findings further underscore the need for closer evaluation of how FFS plans serve this high-need population. The current research findings will help policymakers improve rehabilitation services, quality of care, and outcomes of the current IRF-PPS.

## Supplier

SAS® 9.4

## Sources of Funding

This research was supported by National Institute of Health grants: R01MD017719 (PI: Kumar). The funder played no role in the design, conduct, or reporting of this study.

## Disclosures

The authors have no conflict of interest including finical conflict to declare.

## Data Sharing Statement

The primary data files used for this study are from the Centers for Medicare and Medicaid Services (CMS). According to the Data Use Agreement (DUA), these data files cannot be made publicly available. However, an investigator can obtain these data files directly from the CMS. Dr. Karmarkar had full access to all the data in the study and takes responsibility for the integrity of the data and the accuracy of the data analysis.

## Nonstandard Abbreviations and Acronyms

Inpatient rehabilitation facilities (IRF), Medicare fee-for-service (FFS), Medicare Advantage (MA).

## Data Availability

Due to access restrictions on the CMS data specified in the Data Use Agreement (DUA), we will not be able to make the data available; however, all the data sets used for this study can be obtained directly from the CMS after establishing a DUA.

## Notes

### Competing Interest Statement

The authors have declared no competing interest.

### Author Declarations

Virginia Commonwealth University (VCU)

